# Time restricted eating and sleep in adults with Type 2 Diabetes

**DOI:** 10.1101/2024.07.16.24310541

**Authors:** Vasiliki Pavlou, Shuhao Lin, Sofia Cienfuegos, Mark Ezpeleta, Mary-Claire Runchey, Sarah Corapi, Kelsey Gabel, Faiza Kalam, Shaina J. Alexandria, Alaina P. Vidmar, Krista A. Varady

## Abstract

The aim of this secondary analysis was to compare the effects of time restricted eating (TRE) versus calorie restriction (CR) on sleep in adults with type 2 diabetes (T2D). Adults with T2D (n = 75) were randomized to 1 of 3 interventions for 6-months: 8-h TRE (eating only between 12-8pm daily); CR (25% energy restriction daily); or control. Our results show that TRE has no effect on sleep quality, duration, insomnia severity, or risk of obstructive sleep apnea, relative to CR and controls, in patients with T2D.

## Introduction

Short sleep duration (<5 hours) and poor sleep quality are associated with the risk of developing type 2 diabetes (T2D), and this relationship is thought to be bi-directional.^1^ Individuals with T2D have a higher occurrence of obstructive sleep apnea and can experience disturbances in sleep from diabetes-related nighttime symptoms, such as hypoglycemia, nocturia, or neuropathy.^2,3^ Conversely, laboratory studies show a reduction in insulin sensitivity when sleep duration and quality are decreased in adults with T2D.^4,5^

Weight loss may help improve sleep in those with obesity and T2D. For instance, the Sleep AHEAD trial showed that weight loss of 10.4% in those with T2D improved obstructive sleep apnea symptoms, and even led to sleep apnea remission for some participants, after a 1-year intervention.^6^ In terms of sleep quality and duration, a 2-year weight loss trial in adults with obesity (with and without diabetes), showed improvements in participants who lost more than 5% of their body weight at 6 months, compared to those that lost less than 5%. ^7^

Time restricted eating (TRE) is a weight loss intervention that involves confining food intake to 4 to 10 hours per day and fasting for the remaining hours with energy-free beverages.^8^ TRE results in an unintentional reduction in energy intake of ∼300-500 kcal/d, and weight loss of ∼3-5% over 2-12 months in adults with obesity.^8^ More recently, our group showed that TRE is also effective for weight loss in patients with T2D. ^9^ What remains unknown, however, is whether these changes in body weight have any impact on sleep parameters in this population. Accordingly, we conducted this secondary analysis to compare the effects of 6 months of TRE versus CR and controls on sleep quality, duration, insomnia severity, and risk of obstructive sleep apnea, in adults with T2D. We hypothesized that the TRE group would achieve greater improvements in sleep quality and duration, compared with the CR and control groups, due to greater weight loss by TRE participants.

## METHODS

### Trial participants

This is a secondary analysis of a 6-month randomized, controlled, trial. ^9^ Participants were recruited from the Chicagoland area. Inclusion criteria: 18 to 80 years; previous diagnosis of T2D; HbA1c between 6.5-11.0%; and BMI between 30-50 kg/m^2^. Exclusion criteria: > 4% weight loss or gain in the 3 months prior to study start; eating within less than a 10-h eating window; pregnant or attempting to conceive; history of eating disorders; nightshift work; and currently smoking. The protocol was approved by the Office for the Protection of Research Subjects at the University of Illinois Chicago, and written informed consent was obtained from all participants.

### Intervention groups

Participants were randomized to 1 of 3 groups: TRE, CR or control, by a stratified random sampling procedure based on sex, age, BMI, and HbA1c. TRE participants were instructed to eat ad libitum between 12:00 and 8:00 pm daily and fast from 8:00 to 12:00 pm the following day. Within the 8-h eating window, participants were not restricted on the types or quantities of food consumed. TRE participants were allowed to consume energy-free beverages during the 16-hour fasting period. CR participants were instructed to reduce their energy intake by 25% of their baseline energy needs every day. Total energy expenditure was calculated using the Mifflin equation,^10^ and CR participants were asked to track their food intake daily using an app. Control participants were instructed to maintain their weight by not changing their eating or activity routines.

### Outcome measures

All variables were assessed at baseline and month 6. ^9^ Briefly, body weight was measured at the research center using a digital scale. Fat mass, lean mass, and visceral fat mass were measured by dual-energy x-ray absorptiometry (iDXA, GE). HbA1c levels were analyzed by a commercial lab (MedStar, IL). Time in euglycemic range (i.e., glucose levels between 70-180 mg/dL) was assessed using a continuous glucose monitor (CGM; Dexcom G6 Pro). Physical activity (steps/d) was assessed using a pedometer (Fitbit Alta). Dietary intake was measured using a 7-d food record by the NIH Automated Self-Administered 24-h Dietary assessment tool (ASA-24).^11^

Sleep quality, timing, and duration were measured using the Pittsburgh Sleep Quality Index (PSQI).^12^ The PSQI is a 19-item self-report questionnaire that measures sleep quality in the past month, resulting in a total score of 0-21. Poor sleep quality is indicated by scores above 5. Sleep onset latency was also measured by the PSQI. The sleep latency score ranges from 0-3, with 0 indicating no problem falling asleep, and 3 indicating a severe problem. Insomnia severity was assessed by the Insomnia Severity Index (ISI).^13^ The ISI is a 7-item self-report questionnaire that rates each item by a 5-point Likert scale, resulting in a total score of 0-28. Scores are stratified into the following categories: no clinically significant insomnia (0-7); subclinical insomnia (8-14); moderate severity insomnia (15-21); and severe insomnia (22-28). Risk of obstructive sleep apnea (% occurrences) was estimated using the 10-item self-report Berlin Questionnaire. ^14^ Chronotype was quantified by the Morningness-Eveningness Questionnaire (MEQ), and scores are stratified as follows: definitely morning type (70–86), moderately morning type (59–69), intermediate type (42– 58), moderately evening type (31–41), and definitely evening type (16–30).^15^

### Statistical analysis

Data are shown as mean (95% confidence interval [CI]) unless otherwise noted. We conducted an intention-to-treat analysis, which included data from all 75 participants who underwent randomization. A linear mixed model was used to assess time, group, and time*group effects for each outcome. In each model, time and group effects (and their interaction) were estimated without imposing a linear time trend. A Bonferroni-adjusted two-tailed P value of less than 0.017 was considered statistically significant for pairwise group comparisons of percent change in body weight. P values generated from analyses of secondary outcomes were not adjusted for multiplicity and are considered descriptive. Pearson correlations were performed to assess the relationships between changes in body weight, HbA1c, and sleep measures. All analyses were performed using R software (version 4.3.1).

## RESULTS

### Subject baseline characteristic and dropouts

As previously reported,^9^ a total of 127 participants were screened and 75 were randomized to the TRE group (n = 25), CR group (n = 25) or control group (n = 25). By the end of the 6-month study, the number of completers was n = 69 (TRE: n = 23; CR: n = 22; control: n = 24). Baseline characteristics of the participants were comparable between groups (**Table 1**). Participants in the TRE, CR, and control groups were classified in the intermediate chronotype (i.e., MEQ score between 42–58).

**Table 1.** Baseline characteristics.

| Variables | Time restricted eating (TRE) | Daily calorie restriction (CR) | Control (CON) |
| --- | --- | --- | --- |
| **n** | 25 | 25 | 25 |
| **Age (y)** | 56 (13) | 55 (13) | 54 (11) |
| **Sex (no. female/male)** | 18 / 7 | 17 / 8 | 18 / 7 |
| **Diabetes duration (y)** | 15 (11) | 14 (9) | 12 (9) |
| **Body weight & composition** |  |  |  |
| Body weight (kg) | 105 (25) | 104 (17) | 107 (22) |
| Fat mass (kg) | 43 (10) | 49 (10) | 47 (14) |
| Lean mass (kg) | 54 (13) | 52 (10) | 53 (10) |
| Visceral fat mass (kg) | 1.9 (0.7) | 2.5 (1.0) | 2.2 (0.9) |
| Waist circumference (cm) | 117 (13) | 120 (9) | 121 (15) |
| Body mass index (BMI; kg/m<sup>2</sup>) | 39 (9) | 38 (5) | 39 (7) |
| **Glycemic control** |  |  |  |
| HbA1c (%) | 8.3 (2.0) | 8.1 (1.5) | 7.9 (1.3) |
| Time in euglycemic range (%) | 63 (30) | 60 (30) | 62 (32) |
| **Pittsburgh Sleep Quality Index (PSQI)** |  |  |  |
| Total PSQI sleep quality score | 7.6 (3.2) | 8.2 (3.4) | 9.4 (3.8) |
| Wake time (h:min) | 6:22 (1:31) | 6:41 (1:35) | 6:11 (2:14) |
| Bedtime (h:min) | 23:46 (1:46) | 00:35 (1:17) | 00:17 (1:47) |
| Sleep duration (h:min) | 6:36 (1:00) | 6:06 (2:00) | 5:54 (1:36) |
| Sleep onset latency score | 1.3 (1.1) | 1.3 (1.2) | 1.9 (1.1) |
| **Insomnia severity index (ISI)** |  |  |  |
| Total score | 8.2 (5.2) | 7.4 (5.5) | 9.3 (6.2) |
| **Berlin questionnaire** |  |  |  |
| High risk of obstructive sleep apnea (% occurrences) | 65 | 40 | 79 |
| **Morningness-Eveningness (MEQ)** |  |  |  |
| Total score | 58 (8) | 55 (9) | 54 (10) |

Abbreviations: HbA1c: glycated hemoglobin, ISI: Insomnia severity index, PSQI: Pittsburgh Sleep Quality Index. <sup>a</sup> Continuous variables reported as mean ± SD. Participants at high risk of obstructive sleep apnea reported as % occurrences.

### Body weight, body composition, and glucoregulation

Changes in body weight and glucoregulation are displayed in **Table 2** and **Figure 1**. By month 6, body weight significantly decreased in the TRE group (−3.56% [95% CI, −5.92 to −1.20%]), but not in the CR group (−1.78% [95% CI, −3.67 to 0.11%]), versus controls. Fat mass decreased in the TRE group by month 6 (−2.49 kg [95% CI, −4.41 to −0.58 kg]) but not the CR group (−1.65 kg [95% CI, −3.33 to 0.04 kg]), relative to controls. Waist circumference was reduced in the TRE group (−3.44 cm [95% CI, −5.71 to −1.18 cm]) and CR group (−3.50 cm [95% CI, −5.80 to −1.20 cm]), relative to controls, with no differences between TRE and CR. Lean mass and visceral fat mass did not change significantly between groups. HbA1c levels significantly decreased in the TRE group (−0.91% [95% CI, −1.61 to −0.20%]) and CR group (−0.94% [95% CI, −1.59 to −0.30%]), relative to controls, with no differences between the TRE and CR groups. Time in euglycemic range increased in the CR group by month 6 (18.46% [95% CI, 0.31 to 36.62%]) but not the TRE group (12.59% [95% CI, −5.70 to 30.89%]), relative to controls.

**Figure 1.**
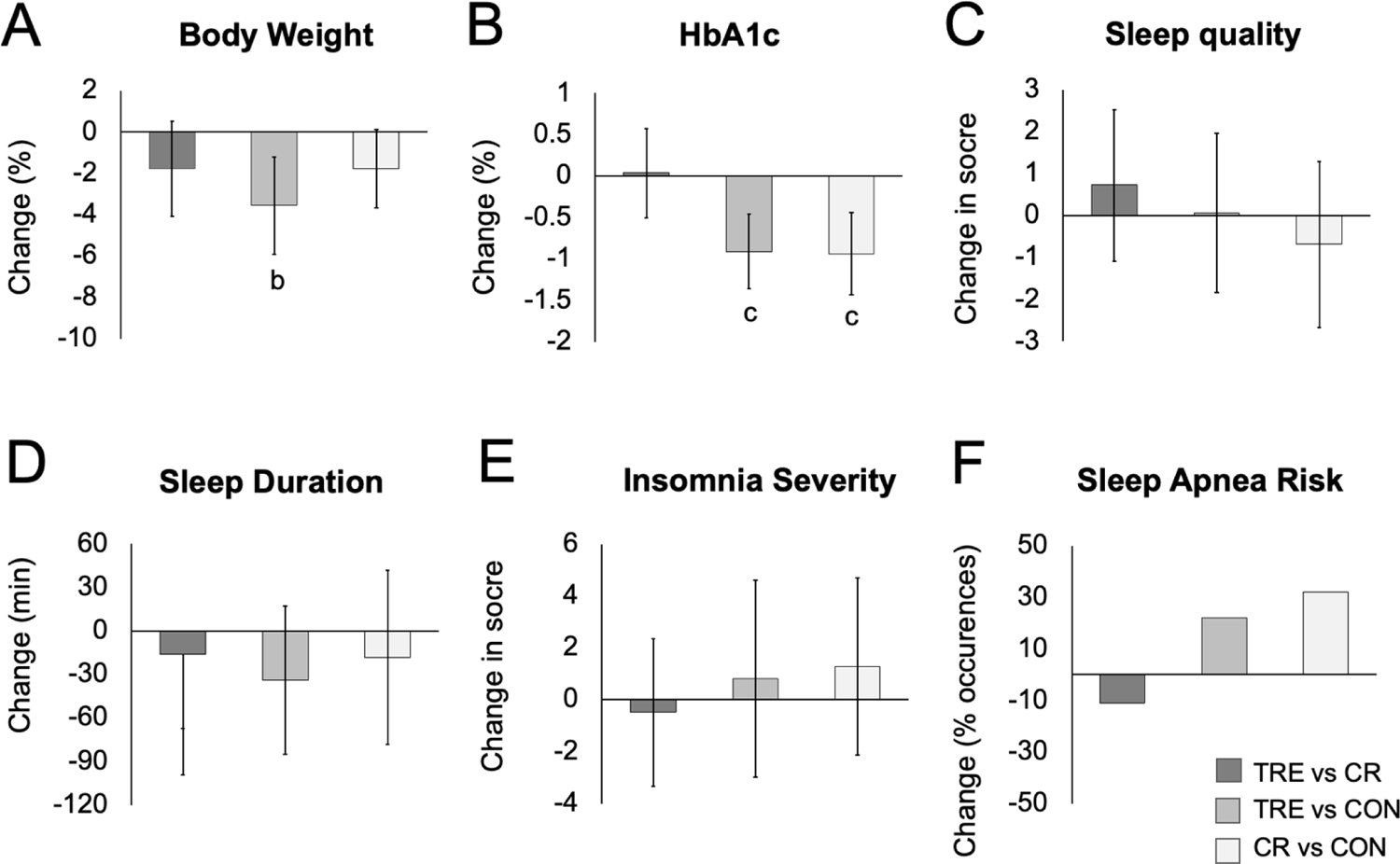
Change in body weight, HbA1c, and sleep parameters between TRE, CR, and control groups Abbreviations: HbA1c: glycated hemoglobin. A. Change in body weight between groups. B. Change in HbA1c between groups. C. Change in sleep quality score between groups. D. Change in sleep duration between groups. E. Change in insomnia severity score between groups. F. Change in risk of obstructive sleep apnea between groups. ^a^ Means were estimated using an intention-to-treat analysis using a linear mixed model with 95% CIs for each parameter from baseline by diet group. ^b^ Indicates statistical significance using Bonferroni-adjusted 2-tailed P < 0.017. ^c^ Indicates statistical significance using P < 0.05.

**Table 2.** Change in body weight, glycemia, and sleep parameters.

| Variables | Change from baseline to month 6 (95% CI) |  |  | Difference between groups by month 6 (95% CI) |  |  |
| --- | --- | --- | --- | --- | --- | --- |
|  | Time restricted eating (TRE) | Daily calorie restriction (CR) | Control (CON) | TRE vs CR | TRE vs CON | CR vs CON |
| **Body weight & composition** |  |  |  |  |  |  |
| Body weight (%) | **-4.28 (-6.26, -2.31) <sup>b</sup>** | **-2.50 (-3.83, -1.18) <sup>b</sup>** | -0.72 (-2.14, 0.69) | -1.78 (-4.09, 0.53) | **-3.56 (-5.92, -1.20) <sup>b</sup>** | -1.78 (-3.67, 0.11) |
| Body weight (kg) | **-4.52 (-6.73, -2.30) <sup>b</sup>** | **-2.63 (-3.99, -1.27) <sup>b</sup>** | -1.07 (-2.70, 0.57) | -1.89 (-4.42, 0.64) | **-3.45 (-6.13, -0.77) <sup>b</sup>** | -1.56 (-3.63, 0.51) |
| Fat mass (kg) | **-2.77 (-4.36, -1.18) <sup>c</sup>** | **-1.92 (-3.20, -0.65) <sup>c</sup>** | -0.28 (-1.46, 0.90) | -0.85 (-2.82, 1.13) | **-2.49 (-4.41, -0.58) <sup>c</sup>** | -1.65 (-3.33, 0.04) |
| Lean mass (kg) | -0.97 (-2.04, 0.10) | -0.46 (-1.15, 0.23) | -0.59 (-1.49, 0.32) | -0.51 (-1.74, 0.72) | -0.38 (-1.74, 0.98) | 0.13 (-0.98, 1.23) |
| Visceral fat mass (kg) | -0.16 (-0.36, 0.05) | **-0.18 (-0.32, -0.04) <sup>c</sup>** | -0.07 (-0.18, 0.04) | 0.02 (-0.21, 0.26) | -0.09 (-0.31, 0.13) | -0.11 (-0.28, 0.06) |
| Waist circumference (cm) | **-3.92 (-5.93, -1.91) <sup>c</sup>** | **-3.97 (-6.03, -1.92) <sup>c</sup>** | -0.48 (-1.66, 0.71) | 0.05 (-2.74, 2.84) | **-3.44 (-5.71, -1.18) <sup>c</sup>** | **-3.50 (-5.80, -1.20) <sup>c</sup>** |
| **Glycemic control** |  |  |  |  |  |  |
| HbA1c (%) | **-0.72 (-1.25, -0.18) <sup>c</sup>** | **-0.75 (-1.20, -0.30) <sup>c</sup>** | 0.19 (-0.30, 0.68) | 0.04 (-0.64, 0.72) | **-0.91 (-1.61, -0.20) <sup>c</sup>** | **-0.94 (-1.59, -0.30) <sup>c</sup>** |
| Time in euglycemic range (%) | 4.78 (-3.52, 13.08) | **10.65 (2.65, 18.65)** | -7.81 (-24.90, 9.27) | -5.87 (-17.03, 5.28) | 12.59 (-5.70, 30.89) | **18.46 (0.31, 36.62)** |
| **Pittsburgh Sleep Quality Index (PSQI)** |  |  |  |  |  |  |
| Total PSQI sleep quality score | -0.60 (-1.84, 0.65) | -1.33 (-2.71, 0.05) | -0.65 (-2.15, 0.85) | 0.73 (-1.07, 2.54) | 0.06 (-1.84, 1.95) | -0.68 (-2.66, 1.30) |
| Wake time (h:min) | -00:05 (-00:35, 00:26) | 00:04 (-00:28, 00:36) | 00:17 (-00:24, 00:59) | -00:08 (-00:52, 00:34) | -00:22 (-01:12, 00:28) | -00:14 (-01:04, 00:37) |
| Bedtime (h:min) | 00:21 (-00:12, 00:54) | 00:14 (-00:27, 00:56) | -00:26 (-00:58, 00:05) | 00:07 (-00:45, 00:58) | **00:47 (00:03, 01:32) <sup>c</sup>** | 00:41 (-00:10, 01:32) |
| Sleep duration (h:min) | -00:20 (-00:50, 00:09) | -00:05 (-00:47, 00:38) | 00:14 (-00:31, 00:58) | -00:16 (-01:07, 00:35) | -00:34 (-01:25, 00:17) | -00:18 (-01:18, 00:42) |
| Sleep onset latency (h:min) | 00:12 (-00:21, 00:44) | -00:12 (-00:42, 00:17) | -00:31 (-01:09, 00:06) | 00:24 (-00:19, 01:07) | 00:43 (-00:05, 01:31) | 00:19 (-00:27, 01:05) |
| **Insomnia severity index (ISI)** |  |  |  |  |  |  |
| Total score | -0.81 (-3.21, 1.60) | -0.31 (-2.02, 1.39) | -1.60 (-4.68, 1.48) | -0.49 (-3.34, 2.36) | 0.80 (-3.00, 4.59) | 1.29 (-2.13, 4.71) |
| **Berlin questionnaire** |  |  |  |  |  |  |
| Risk of obstructive sleep apnea (%) | -13 (-28, 3) | -2 (-29, 25) | -34 (-56, -12) | -11 (-41, 20) | 22 (-5, 48) | 32 (-1, 66) |

Abbreviations: HbA1c: glycated hemoglobin, ISI: Insomnia severity index, PSQI: Pittsburgh Sleep Quality Index.
<sup>a</sup> Means were estimated using an intention-to-treat analysis using a linear mixed model with 95% CIs for each parameter from baseline by diet group.
<sup>b</sup> Indicates statistical significance using Bonferroni-adjusted 2-tailed P < 0.017.
<sup>c</sup> Indicates statistical significance using P < 0.05.

### Energy intake and physical activity

The mean (SD) energy deficit was −313 (SD 509) kcal/d in the TRE group, −197 (SD 426) kcal/d in the CR group, and −16 (439) kcal/d in the control group over 6 months. Diet quality and physical activity did not differ over time or between groups, as reported previously. ^9^

### Sleep measures

Changes in sleep measures are reported in **Table 2** and **Figure 1**. At baseline, mean (SD) sleep quality was poor (PSQI scores >5) in the TRE (7.6 ± 3.2), CR (8.2 ± 3.4), and control groups (9.4 ± 3.8). The PSQI sleep quality score did not change significantly between groups by month 6. At baseline, mean (SD) sleep duration was shorter than the recommended 7 h minimum per night^16^ in TRE (6:36 ± 1:00 h:min), CR (6:06 ± 2:00 h:min), and control group (5:54 ± 1:36 h:min). Sleep duration did not change significantly between groups by month 6. Wake time and sleep onset latency also did not change significantly between groups by the end of the study. As for bedtime, the TRE group was going to sleep later by month 6 (00:47 h:min [95% CI, 00:03 to 01:32 h:min]), relative to controls only.

At baseline, the insomnia severity score (SD) was 8.2 (5.2) for the TRE group and 9.3 (6.2) for the control group, indicating subclinical insomnia (ISI score 8-14), and 7.4 (5.5) in the CR group, indicating no clinical insomnia (ISI score 0-7). Insomnia severity scores did not change significantly between groups by month 6. At baseline, the risk for obstructive sleep apnea was present in 65% of TRE participants, 40% of CR participants, 79% of control participants. Risk of obstructive sleep apnea did not change significantly between groups by the end of the study. Changes in body weight and HbA1c were not related to any sleep measure in any group.

## Discussion

To our knowledge this is the first study to compare the effects of TRE to CR on sleep outcomes in adults with T2D. Our results show that TRE produced significant reductions in body weight and HbA1c levels after 6 months, but no changes in sleep quality, duration, insomnia severity, or risk of obstructive sleep apnea, versus CR and controls.

Sleep quality and duration (measured by PSQI) remained unchanged in all groups by the end of the study. This finding is similar to what was reported in other TRE studies.^17–19^ For instance, Parr et al.^19^ reported no change in body weight and sleep quality or duration after one month of 9-h TRE (10am-7pm) in adults with obesity and T2D. Gabel et al.^18^ showed that 3 months of 8-h TRE (10am-6pm) did not alter sleep quality or duration in adults with obesity even with a 2.6% weight loss.

Similarly, Cienfuegos et al.^17^ reported no change in sleep quality or duration after 2 months of 4-h TRE (3-7pm) and 6-h TRE (1-7pm) versus controls in adults with obesity with a 3.2% weight loss. It is possible that sleep quality and duration were not improved in any of these TRE trials because the degree of weight loss fell short of being clinically significant (>5% weight loss from baseline). In trials that reported greater weight loss, such as the DiRECT trial^20^ (8.8% weight loss), sleep quality and duration increased over 24 months in adults with obesity and T2D. Evidence suggests that weight loss may improve sleep quality and duration by reducing sleep fragmentation and alleviating sleep-disordered breathing.^7^

Insomnia severity was also assessed. At the beginning of the study, participants in the TRE and control groups portrayed sub-clinical insomnia, while subjects in the CR group displayed no clinically significant insomnia. By the end of the trial, no changes in insomnia severity were noted in the intervention groups versus controls. This finding is not surprising as our subjects did not portray clinically significant insomnia at the onset of treatment; therefore, it would be unlikely for this sleep metric to improve. This lack of effect on insomnia severity has also been observed other TRE trials that observed a similar degree of weight loss (∼3%).^17,18^

Obstructive sleep apnea is highly prevalent in those with T2D.^21^ In the present trial, 65-79% of subjects in the TRE and control groups were at high risk of sleep apnea, while 40% of participants in the CR group were at high risk. After 6 months, no significant changes in the risk of sleep apnea were noted. However, it is possible that our interventions did not achieve enough weight reduction to improve this sleep metric. Findings suggest that at least 10% weight loss may be necessary to decrease the risk of obstructive sleep apnea in people with obesity.^22^ Indeed, the Sleep AHEAD study showed an improvement in obstructive sleep apnea (measured by apnea-hypopnea index) with a 10% weight loss after 1 year of intensive lifestyle intervention, in 264 participants with obesity and T2D. ^6^

This study has several limitations. First, our sample size was small (n = 75) and our power calculation was based solely on changes in body weight. Thus, it is likely that our study was not powered adequately to identify significant changes in sleep parameters. Second, self-report was used to assess all sleep outcomes. This study would have benefitted from the use of wrist actigraphy to provide more objective assessments of rest and activity patterns. Third, participants were allowed to consume caffeinated beverages during their fasting window. As such, some participants may have consumed caffeine late into the evening, which may have impacted their sleep. Fourth, the study may have been too short (6-months) to produce any meaningful changes in body weight and sleep parameters.

In summary, the weight loss induced by 6-months of TRE does not improve sleep quality, duration, insomnia severity or risk of obstructive sleep apnea in individuals with obesity and T2D, relative to CR and controls. However, these findings will need to be confirmed by a well-powered randomized controlled trial specifically designed to assess the impact of TRE on sleep in this population group.

## Data Availability

All data produced in the present work are contained in the manuscript

## ACKNOWLEDGEMENTS

Conflict of interest:

KAV reports receiving author fees from Pan MacMillan Publishing for the book, “The Fastest Diet.” The other authors declare no conflict of interest.

## Author contributions

VP designed the research, conducted the clinical trial, analyzed the data, and wrote the manuscript; SL, SC, ME, MCR, SC, KG, and FK assisted with the conduction of the clinical trial; SJA performed the statistical analysis, APV served as the study endocrinologist; KAV designed the research, and wrote the manuscript.

## Trial registration

ClinicalTrials.gov, NCT05225337.

